# The UK Soft Drinks Industry Levy and childhood hospital admissions for asthma in England

**DOI:** 10.1101/2023.11.06.23298141

**Authors:** Nina T Rogers, Steven Cummins, Catrin P Jones, Oliver T Mytton, Chrissy H Roberts, Seif O. Shaheen, Syed Ahmar Shah, Aziz Sheikh, Martin White, Jean Adams

## Abstract

Sugar sweetened beverage consumption has been suggested as a risk factor for asthma symptoms in children. We examined whether the UK Soft Drinks Industry Levy (SDIL), announced in March 2016 and implemented in April 2018, was associated with changes in National Health Service hospital admission rates for asthma in children.

We conducted interrupted time series analyses (2012 - 2020) to measure changes in monthly incidence rates of hospital admissions for asthma. Sub-analysis was by age-group (5-9, 10-14, 15-18 years) and neighbourhood deprivation quintiles. Estimated changes were relative to counterfactual scenarios where SDIL was neither announced, nor implemented. Compared to the counterfactual scenario there was an overall relative reduction in incidence rates of hospital admissions of 20.9% (95%CI: 29.6-12.2), 22 months post-SDIL. Reductions were similar across age-groups and deprivation quintiles with relative reductions of 24.3% (95%CI: 29.6-12.2) in children aged 10-14 years, and reductions of 23.2% (95%CI 33.4-13.1) in children living in the middle deprivation quintile. These findings provide evidence that the implementation of a tax intended to reduce childhood obesity in the UK preceded a potentially significant collateral public health benefit in the form of reduced hospital admissions for childhood asthma.

## INTRODUCTION

The UK Scientific Advisory Committee on Nutrition (SACN) recommends that consumption of free sugar should be below 5% of total energy intake. In the UK, consumption across all age groups is at least twice this, and three times greater in adolescents aged 11-18 years^1^. Sugar sweetened beverages (SSBs) are a major source of free sugar in the UK diet^2^ and high intake has consistently been found to be associated with risk of non-communicable diseases, including obesity, diabetes, and cardiovascular disease^3^. There is also increasing evidence of a link between SSB consumption and incidence of asthma^4^, one of the most common diseases in childhood and for which the UK has the highest mortality rates in Europe^5^.

A recent meta-analysis based on one cohort study and 11 cross-sectional studies found an increased odds of asthma prevalence when comparing the highest versus lowest consumers of SSBs^6^. Cross-sectional studies have reported associations^7 8^, including dose-response relationships^9^, between consumption of SSBs and prevalence of asthma in children. Furthermore, a high percentage of energy from sugars in SSBs was associated with asthma traits in children in the second year of life^10^. A birth cohort study in the US reported that high intake of fructose-containing beverages in early life (∼3 years old) was associated with an increased risk of ever having asthma (doctor-diagnosed and including taking asthma medication or reporting wheezing in last 12 months) by mid-childhood (∼8 years old)^11^.

While several studies have examined associations between SSB consumption and prevalence or incidence of asthma, we are unaware of any quasi-experimental studies that have investigated the longitudinal impact of changes in sugar content of drinks on asthma outcomes. In March 2016, the UK government announced that a two-tier Soft Drink Industry Levy (SDIL) on drinks manufacturers, designed to incentivise them to reformulate their drinks, would come into force in April 2018^12^, presenting a unique opportunity for a ‘natural experiment’ to examine potential health impacts of the SDIL.

Evidence suggests that the proportion of soft drinks available in the UK with <5g of sugar/100 ml fell from 49% before the announcement (i.e., 2015) to 15% one year following implementation of the SDIL (i.e., 2019)^13^. Household purchasing of sugar from soft drinks also fell by 8g/household/week, with associated health benefits in terms of reductions in obesity prevalence^14^ and carious tooth extractions^15^ in children living in England.

Here, we used National Health Service (NHS) hospital episode data from England, and interrupted time series methodology, to compare trends in the observed incidence of childhood hospital admissions for asthma with a modelled counterfactual scenario where SDIL was not announced or implemented, 22 months after the SDIL came into force (overall, by age-group and by area-based deprivation

## RESULTS AND DISCUSSION

The mean incidence rates of hospital admissions for asthma in children during the pre-announcement and post-announcement period are described in Table 1. This reveals large inequalities, with nearly three times as many children from the most deprived areas attending hospital in the pre-announcement period, equivalent to 26.4/100,000 persons/month (p/m), than in the least deprived areas (9.3/100,000 p/m). Incidence rates for asthma hospital admissions were higher for younger children with those aged 5-9 having around double the rate of children aged 15-18 years.

**Table 1.** Mean incidence rates (standard deviation) in the pre^1^ and post^2^ announcement periods of the UK SDIL and change* in hospital admissions per 100,000 population per month for asthma, overall and by age groups and IMD quintiles.

|  | Mean Incidence of asthma hospital admissions / 100,000 p/m |  | Change in asthma admissions compared to the counterfactual scenario |  |
| --- | --- | --- | --- | --- |
|  | Pre-announcement<sup>1</sup> | Post-announcement<sup>2</sup> | Absolute Change | Relative Change (%) |
| Total population (age 5-18) | 15.8 (4.0) | 16.5(3.9) | **-4.0(-2.4, -5.7)** | **-20.9( -29.6, -12.2)** |
| Age-groups |  |  |  |  |
| 5-9 | 22.0(7.1) | 21.3(6.7) | **-4.3(-6.9, -1.7)** | **-18.6(-30.0, -7.2)** |
| 10-14 | 14.9(3.1) | 15.4(2.8) | **-4.7(-6.2, -3.2)** | **-24.3(-32.1, -16.5)** |
| 15-18 | 9.0(2.0) | 11.3(2.0) | **-2.1(-2.6, -1.5)** | **-15.6(-19.7, -11.5)** |
| IMD fifths |  |  |  |  |
| IMD1:Most deprived | 26.4 (7.03) | 28.6 (7.0) | **-4.8 (-7.4, -2.25)** | **-15.5 (-23.7, -7.19)** |
| IMD2 | 17.6 (4.4) | 18.7 (4.5) | **-4.9 (-6.94, -2.8)** | **-22.1 (-31.6, -12.6)** |
| IMD3 | 13.3(3.6) | 13.6 (3.3) | **-3.8 (-5.47, -2.1)** | **-23.2 (-33.4, -13.1)** |
| IMD4 | 11.6 (2.9) | 11.4 (2.8) | **-3.1 (-4.39, -1.7)** | **-23.2 (-33.3, -13.0)** |
| IMD5: least deprived | 9.3(2.6) | 9.8 (2.4) | **-3.4 (-4.41, -2.3)** | **-26.4 (-34.6, -18.1)** |

\*Change in incidence rate of hospital admissions is compared to a counterfactual scenario of the UK SDIL not being announced or implemented.

### Changes in asthma hospital admissions in relation to the SDIL

In children aged 5-18 years there was an overall absolute reduction in hospital admissions for asthma of 4.0 (2.4, 5.7)/100,000 p/m, or a relative reduction of 20.9% (95%CI: 29.6, 12.2), compared to the counterfactual scenario of no SDIL announcement and no implementation (see Table 1 and Figure 1). Upward trends were observed in overall asthma admissions until a few months after the SDIL announcement when a downward trend was observed (Figure 1). Dips in admissions appeared to occur in April and August each year (coincident with school holidays). Large spikes were observed in early autumn each year, especially September, a time previously reported to be associated with a sharp peak in childhood asthma hospitalisations^16^. Possible explanations include the start of the new school year, when there is a high exposure to respiratory viruses, and allergens in schools (including dust mites), increased stress from starting school and children have gotten out of the routine of taking their preventor inhalers following the summer months^16–18^.

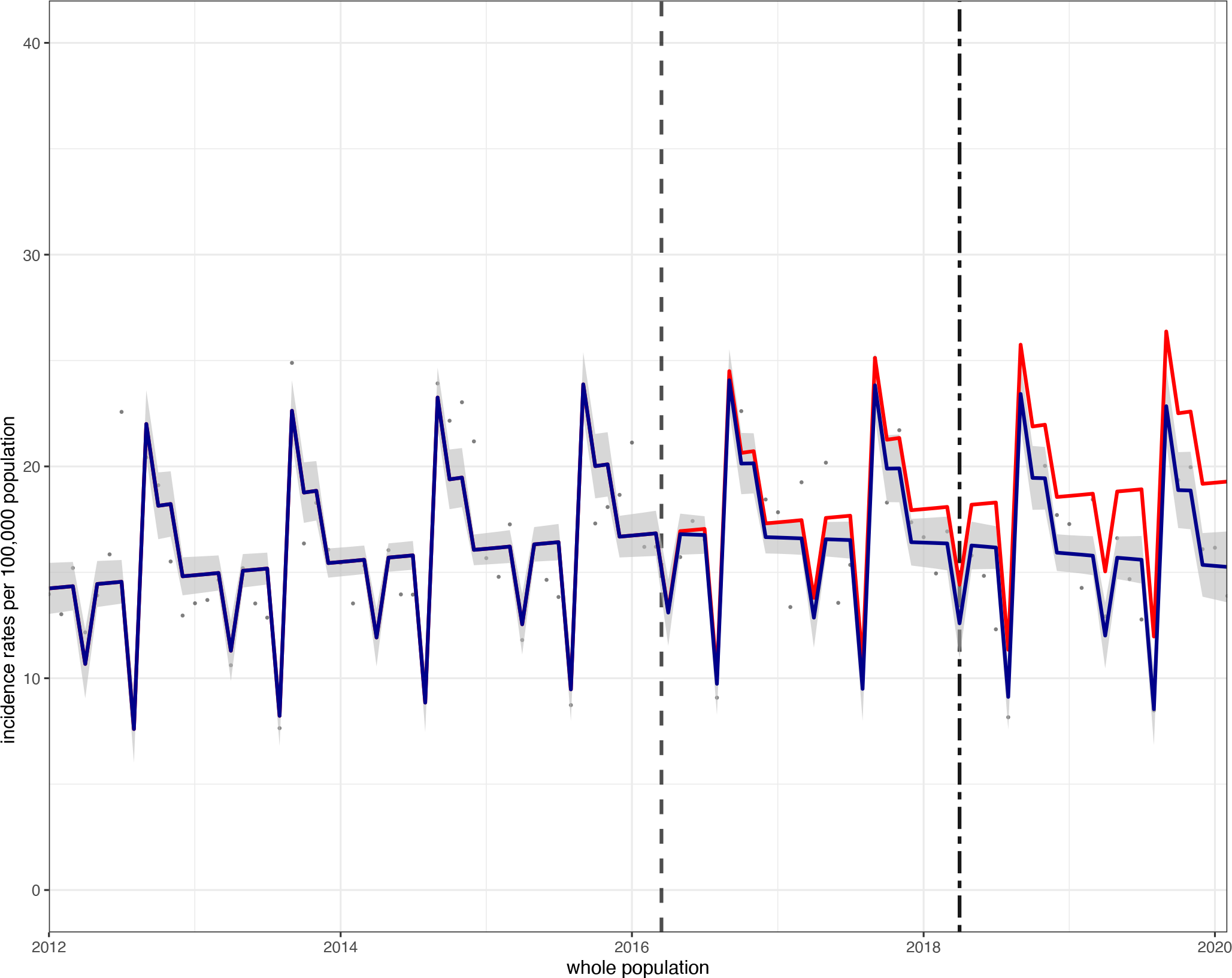

In each age group, there were also upward trends in incidence of hospital admission rates for asthma from the start of the study-period, but in all cases compared to the counterfactual there were significant reductions, 22 months following the implementation of the SDIL (Figure 2). In age-groups 5-9 and 10-14 years, there were relative reductions of 18.6% (95%CI: 30.0, 7.2) and 24.3% (95%CI: 32.1, 16.5), respectively and visualisations suggested a reversal of the upward trend occurring after the SDIL announcement. For adolescents aged 15-18 years, there was a relative reduction of 15.6% (95%CI: 19.7, 11.5) with a flattening out, but not a reversal of the pre-announcement upward trend in hospital admissions.

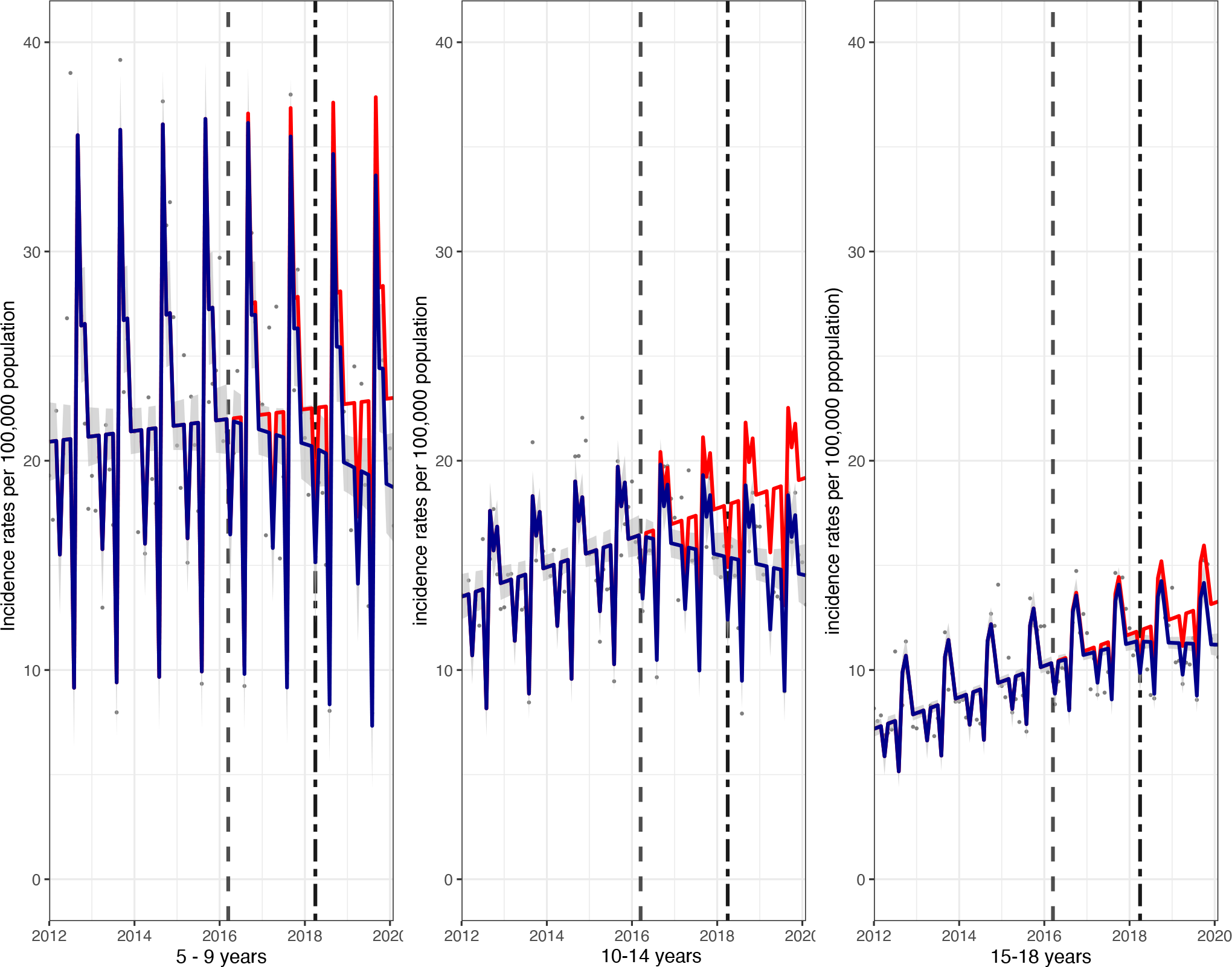

Hospital admissions for childhood asthma dropped across all deprivation groups. Absolute reductions were 4.8 (7.4, 2.3)/100,000 p/m and 3.4 (4.4, 2.3)/100,000 p/m, in the most and least deprived quintiles, respectively. Comparable relative reductions were 15.5% (95%CI: 23.7, 7.2) and 26.4% (96%CI: 34.6, 18.1), respectively (Figure 3). Absolute reductions were relatively consistent across the different IMD quintiles, however, some evidence for a trend in higher relative reductions in less deprived areas were observed (Table 1).

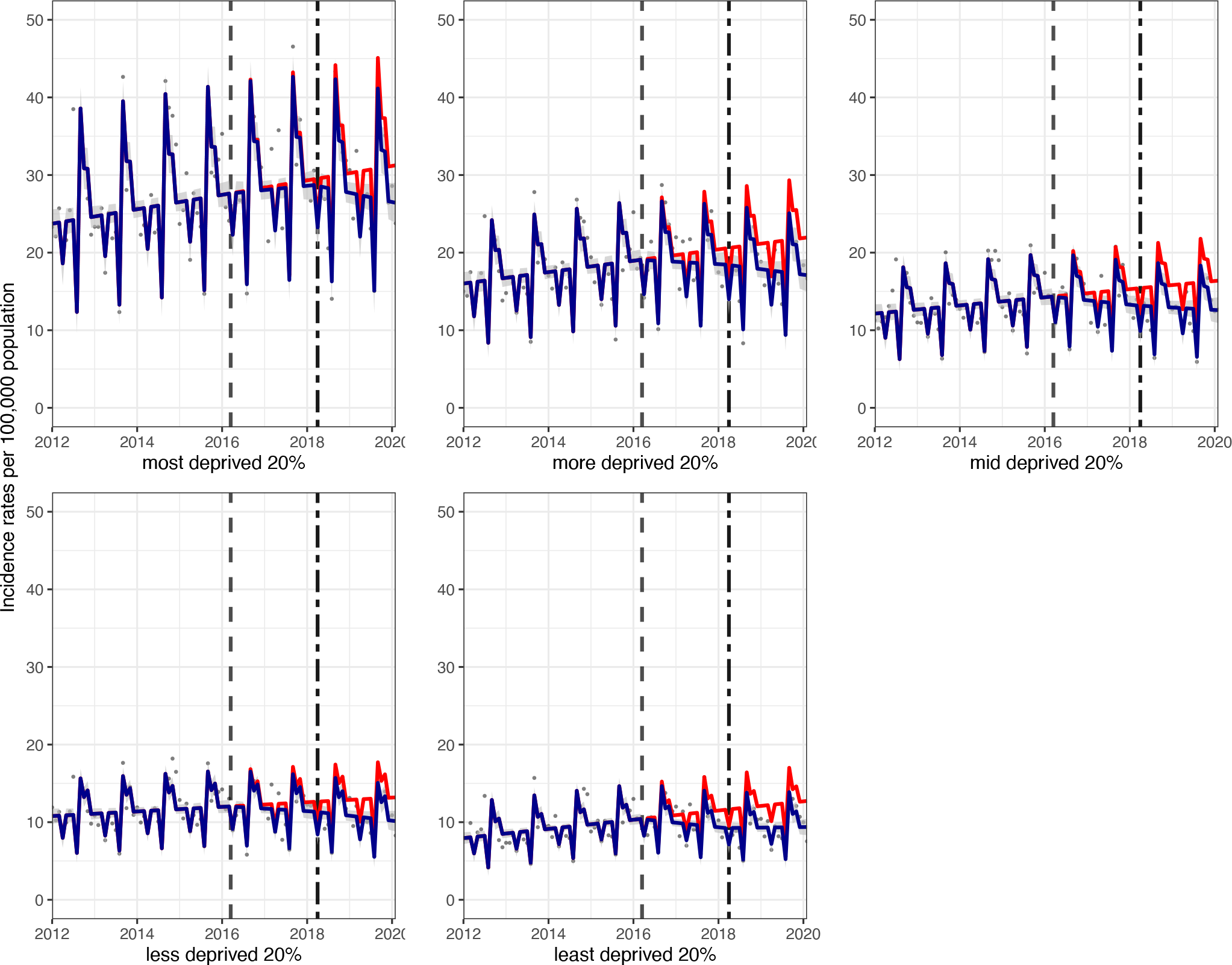

## Discussion

This study provides the first natural experimental evidence to estimate the effects of SSB taxes on asthma outcomes in children. The SDIL was associated with an overall reduction in hospital admissions for asthma in children aged 5-18 years of 21%, 22 months after implementation. Reductions were seen across all areas of deprivation and age groups.

Our findings are consistent with previous studies that suggest an association between SSB consumption and asthma outcomes^7,9–11^, but arguably the quasi-experimental design of our study provides stronger evidence for a causal relationship. Sugar intake has been linked to the development of asthma, with maternal intake of free sugar found to be a risk factor for asthma and atopy in offspring^19^. Ecological evidence suggests an association between per capita consumption of sugar during the perinatal period and severe asthma in 6-7 year old children^20^.

Possible explanations for a potential effect of SSB consumption on asthma include obesity (linked to SSB consumption), which is a risk factor for asthma, with factors such as systemic inflammation and nutrition potentially having a moderating effect^21^. On the other hand, most studies have found that that the SSB consumption-asthma link is independent of body mass index (BMI) or obesity status^7,9–11^. It is also noteworthy that the SDIL was not associated with a significant reduction in the prevalence of obesity in all groups of children^14^. Secondly, preservatives in SSBs have been suggested as a precipitating factor in asthma attacks ^22^ however studies have not found a link between diet soft drinks, which also contain the same preservatives^7^. Thirdly, high fructose: glucose ratios in SSBs have been implicated in asthma risk with suggestions that reducing consumption may lower asthma risk^4^. Inflammation caused by advanced glycation end products, from fructose, may be an important mechanism for the link between fructose consumption and asthma in children. Alternatively, fructose also causes generation of uric acid, and experimental evidence in mice suggests that uric acid promotes Th2 cell-dependent allergic inflammation^23^. This may have important policy implications. For example, 100% fruit juice is a significant contributor of free sugars in the diets of school-age children, contains high levels of free fructose, and has been associated with an increased risk of asthma in children^7 8^, but is currently exempt from the SDIL.

Strengths of our study include using routinely collected hospital admission data on asthma in all children attending an NHS hospital which are therefore not influenced by response bias. We also included a counterfactual scenario in our methodology. One consideration in ITS analysis is the vulnerability to time-varying confounding by other interventions (or treatments) that may have been implemented at around the same time as the SDIL, and that are difficult to account for. We are unaware of any such intervention, although acknowledge that in October 2015 a national law that prohibited smoking inside private-vehicles if children were inside came into force^24^. However no association between introduction of this smoke-free vehicle legislation and childhood tobacco smoke exposure or respiratory health has been found, suggesting it is unlikely to be a source of time varying confounding ^25^.

Our findings link the SDIL to absolute reductions that are of a similar magnitude across deprivation levels. Lower relative reductions were observed in more deprived areas because incidence rates for asthma hospitalisation are higher in children living in more deprived areas. Deprivation is linked to exposure to tobacco smoke, pollution, psychosocial stress and poor management of asthma, and is therefore a risk factor for asthma ^26^.Our findings indicate that the SDIL alone will not reduce inequalities in asthma hospitalisations and should be implemented alongside other strategies to reduce inequalities in asthma.

## METHODS

### Data source

National Health Service (NHS) hospital admissions for asthma (International Classification of Diseases; ICD-10 code: J45) in children aged 5-18y were identified using Hospital Episodes Statistics (HES) data. Data was analysed (1) overall, (2) by age-groups 5-9, 10-14 and 15-18 years and (3) by Index of Multiple Deprivation (IMD) quintile of the Lower Super Output Area (LSOA) of residence ^27^. We did not analyse admissions attributed to asthma under five years of age because it is difficult to diagnose asthma at this age when preschool wheeze, a different phenotype, predominates^28^. In HES, age of the patient was calculated from the birth date and episode start date and the 2010 version of the IMD was used to rank LSOAs according to deprivation and assigned into fifths based on the distribution of all LSOAs in England. The study period was from January 2012 (study month 01) to February 2020 (study month 98) and included the time of the SDIL announcement (March 2016; study month 51) and date when the SDIL came into force (April 2018; study month 76). The study was curtailed a month prior to national lockdown of the COVID-19 pandemic to avoid the possibility that admissions after this date may have been related to COVID-related wheezing in children. Under the SDIL, manufacturers of soft drinks containing ≥ 8g of sugar/100ml and those with ≥5 to <8g of sugar / 100ml were subject to a levy of £0.24/litre and £0.18/litre, respectively. Soft drinks containing <5g/100ml sugar, 100% fruit juice and milk and milk based drinks and drinks with ≥ 1.2% alcohol by volume were levy-exempt.

### Statistical analysis

Interrupted time series (ITS) analyses were conducted to examine changes in incidence rates of childhood hospital admissions for asthma relative to a counterfactual scenario where the SDIL was not announced or implemented. Polynomial regression was used to estimate groupwise (e.g. age 5-9 years) population sizes in each study month. The groupwise number of admissions was divided by the respective estimated population size and multiplied by 100,000 to give an incidence rate of hospital admissions per 100,000 population^29^. Time series models were based on generalised least squares (GLS).

Calendar months were tested to determine variation in monthly admissions and GLS models included any month that was associated with significant changes in incidence of hospital admissions. Models were adjusted for September, October, November, April and August, where there were statistically significant changes in incidence rates of hospital admissions.

Counterfactual scenarios were modelled from pre-announcement trend data (study months 01–51). Confidence intervals for calculated differences were estimated from standard errors that were generated from the delta method^30^.

Autocorrelation was examined statistically using Durbin–Watson tests and visually using graphical plots of autocorrelation and partial autocorrelation. For individual models we used an autocorrelation-moving average (ARIMA) correlation structure, with the order (p) and moving average (q) parameters specifically selected to minimise the Akaike information criterion (AIC) in individual models.

Statistical analyses were conducted in R version 4.1.0.

## Data Availability

Data was acquired through a data sharing agreement with NHS digital for which conditions
of use apply. Requests for data must be made directly to NHS digital and cannot be granted
by the authors.

